# On the Generation of Medical Dialogues for COVID-19

**DOI:** 10.1101/2020.05.08.20095810

**Authors:** Wenmian Yang, Guangtao Zeng, Bowen Tan, Zeqian Ju, Subrato Chakravorty, Xuehai He, Shu Chen, Xingyi Yang, Qingyang Wu, Zhou Yu, Eric Xing, Pengtao Xie

**Affiliations:** UC San Diego; CMU; UC Davis

**Author notes:** Equal contribution (Wenmian Yang, Guangtao Zeng, Bowen Tan). The work was done during internship at UCSD.

## Abstract

Under the pandemic of COVID-19, people experiencing COVID19-related symptoms or exposed to risk factors have a pressing need to consult doctors. Due to hospital closure, a lot of consulting services have been moved online. Because of the shortage of medical professionals, many people cannot receive online consultations timely. To address this problem, we aim to develop a medical dialogue system that can provide COVID19-related consultations. We collected two dialogue datasets – CovidDialog – (in English and Chinese respectively) containing conversations between doctors and patients about COVID-19. On these two datasets, we train several dialogue generation models based on Transformer, GPT, and BERT-GPT. Since the two COVID-19 dialogue datasets are small in size, which bear high risk of overfitting, we leverage transfer learning to mitigate data deficiency. Specifically, we take the pretrained models of Transformer, GPT, and BERT-GPT on dialog datasets and other large-scale texts, then finetune them on our CovidDialog datasets. Experiments demonstrate that these approaches are promising in generating meaningful medical dialogues about COVID-19. But more advanced approaches are needed to build a fully useful dialogue system that can offer accurate COVID-related consultations. The data and code are available at https://github.com/UCSD-AI4H/COVID-Dialogue

## 1. Introduction

As of May 8th in 2020, the COVID-19 pandemic has killed 272,778 people out of 3,910,738 infected cases. People who are experiencing symptoms (e.g., fever, cough) similar to those of COVID-19 or were exposed to risk factors such as close contact with infected cases have a pressing need to consult doctors, largely because of the panic over this unknown new disease. However, under the pandemic situation, coming to hospitals is dangerous and has high risk of suffering cross-infection. Cross-infection refers to the fact that many people visiting hospitals at the same time and infected individuals will spread coronavirus to healthy ones. To prevent spreading of the coronavirus, many non-urgent clinics and hospitals have been closed physically and encourage people to consult doctors through telemedicine services (e.g., phone calls, video conferencing). However, medical professionals are highly occupied by taking care of the infected patients and have very thin bandwidth to deal with the surging requests of consultations related to COVID-19. As a result, many people could not receive timely advice for effectively dealing with their medical conditions.

To address the large imbalance between the surging need of consultations from citizens and the severe shortage of medical professionals available to provide online consultation services, it is highly valuable to develop intelligent dialogue systems which act as “virtual doctors” to provide COVID-related consultations to people. These “virtual doctors” can greatly ease the burden of human doctors and timely address the concerns of the public.

To facilitate the research and development of COVID19-targeted dialogue systems, we build two medical dialogue datasets that contain conversations between doctors and patients, about COVID-19 and other pneumonia: (1) an English dataset containing 603 consultations, 1232 utterances, and 90664 tokens (English words); (2) a Chinese dataset containing 1088 consultations, 9494 utterances, and 406550 tokens (Chinese characters).

On these two datasets, we train several dialogue generation models based on Transformer (Vaswani et al., 2017), GPT (Radford et al., a; Zhang et al., 2019), and BERT-GPT (Wu et al., 2019; Lewis et al., 2019). Transformer is an encoder and decoder architecture which takes the conversation history as inputs and generates the response. Self-attention is used to capture the long-range dependency among tokens. GPT is a language model based on the Transformer decoder. When generating a response, GPT predicts the next token using its context including the already decoded tokens in this response and the conversation history. BERT-GPT is an encoder-decoder architecture as well where the pretrained BERT (Devlin et al., 2018) is used to encode the conversation history and GPT is used to decode the response. The small size of CovidDialog datasets incurs high risk of overfitting, if directly training the large-sized neural models on CovidDialog. To alleviate this risk, we take the pretrained weights of these models on large-scale dialogue dataset and other corpus and finetune the weights on CovidDialog. Experiments demonstrate that the models trained on CovidDialog datasets are promising in generating clinically meaningful consultations about COVID-19. The datasets and code are publicly available at https://github.com/UCSD-AI4H/COVID-Dialogue

The rest of the papers are organized as follows. Section 2 and 3 present the datasets and methods. Section 4 gives experimental results. Section 5 reviews related works and Section 6 concludes the paper.

## 2. Dataset

In this section, we present two collected datasets – CovidDialog-English and CovidDialog-Chinese – which contain medical conversations between patients and doctors about COVID-19 and other related pneumonia. The statistics of these two datasets are summarized in Table 1.

**Table 1:**
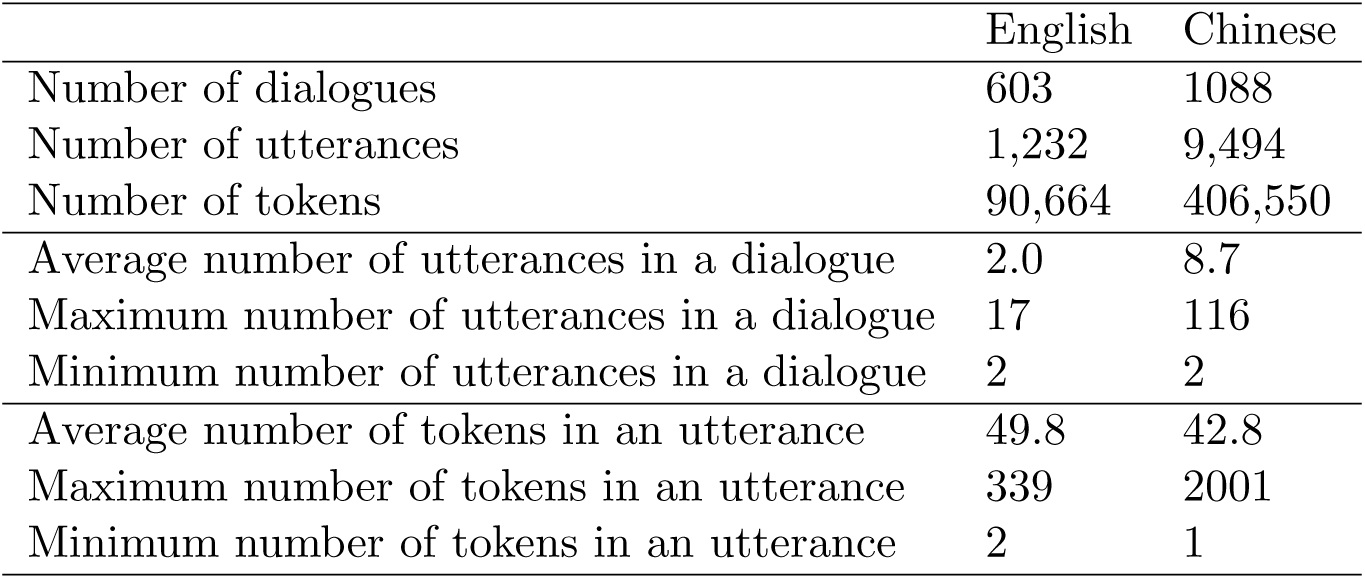
Statistics of the English and Chinese dialogue datasets about COVID-19.

### 2.1. The English Dataset

The CovidDialog-English dataset contains 603 English consultations about COVID-19 and other related pneumonia, having 1,232 utterances. The number of tokens (English words) is 90,664. The average, maximum, and minimum number of utterances in a conversation is 2.0, 17, and 2 respectively. The average, maximum, and minimum number of tokens in an utterance is 49.8, 339, and 2 respectively. Each consultation starts with a short description of the medical conditions of a patient, followed by the conversation between the patient and a doctor. Figure 1 shows an example. The original dialogues are crawled from online healthcare forums, including icliniq.com^1^, healthcaremagic.com^2^, and healthtap.com^3^.

**Figure 1:**
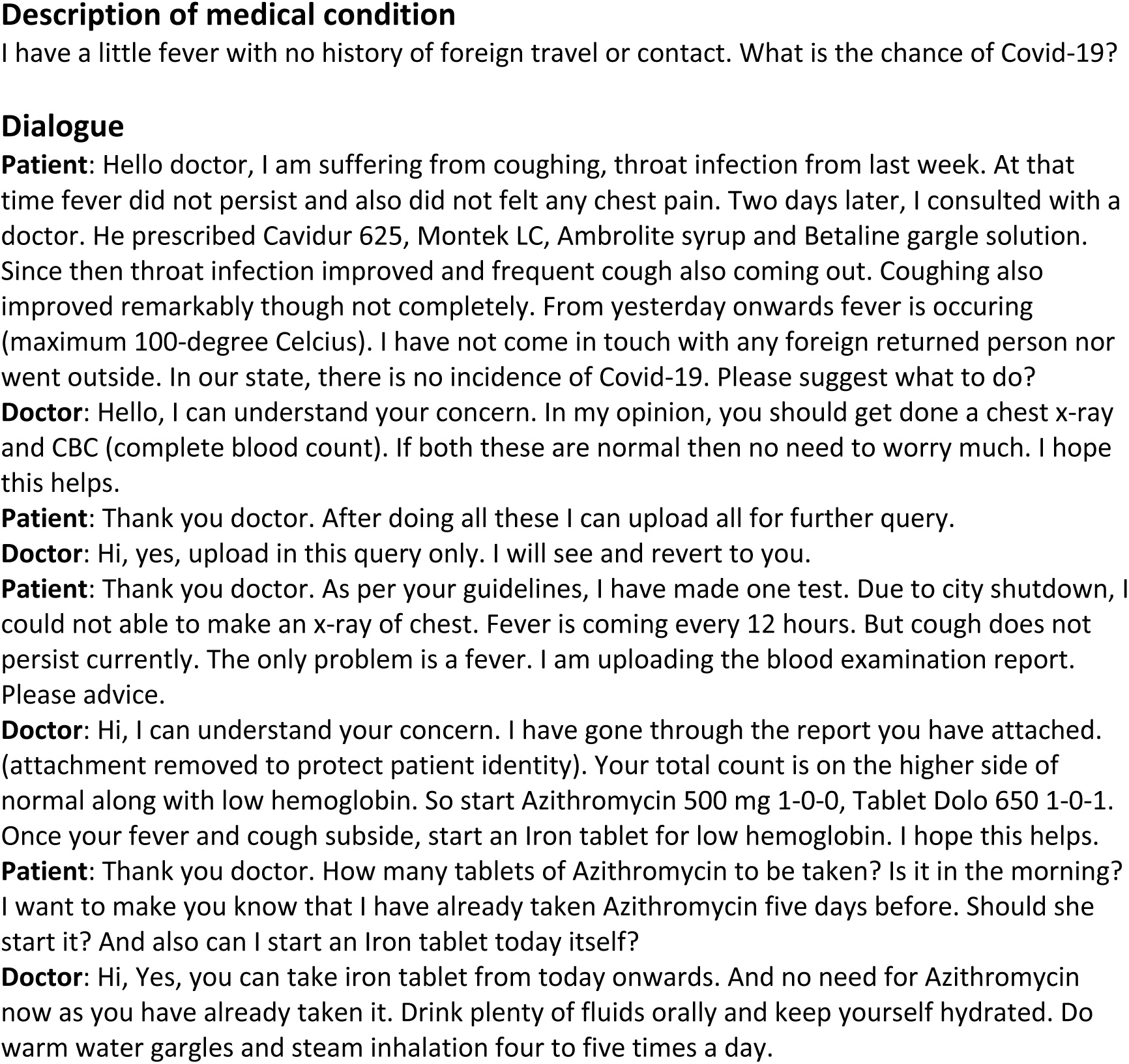
An exemplar consultation in the CovidDialog-English dataset. It consists of a brief description of the patient’s medical conditions and the conversation between the patient and a doctor.

### 2.2. The Chinese Dataset

The CovidDialog-Chinese dataset contains 1,088 Chinese consultations about COVID-19 and other related pneumonia, having 9,494 utterances. In this work, we develop models directly on Chinese characters without performing word segmentation. Each Chinese character in the text is treated as a token. The total number of tokens in the dataset is 406,550. The average, maximum, and minimum number of utterances in a conversation is 8.7, 116, and 2 respectively. The average, maximum, and minimum number of tokens in an utterance is 42.8, 2001, and 1 respectively. Each consultation consists of three parts: (1) description of patient’s medical condition and history; (2) conversation between patient and doctor; (3) (optional) diagnosis and treatment suggestions given by the doctor. In the description of the patient’s medical condition and history, the following fields are included: present disease, detailed description of present disease, what help is needed from the doctor, how long the disease has been, medications, allergies, and past diseases. This description is used as the first utterance from the patient. Figure 2 shows an exemplar consultation. The data is crawled from haodf.com^4^, which is an online platform of healthcare services, including medical consultation, scheduling appointments with doctors, etc. Duplicated and incomplete dialogues were removed.

**Figure 2:**
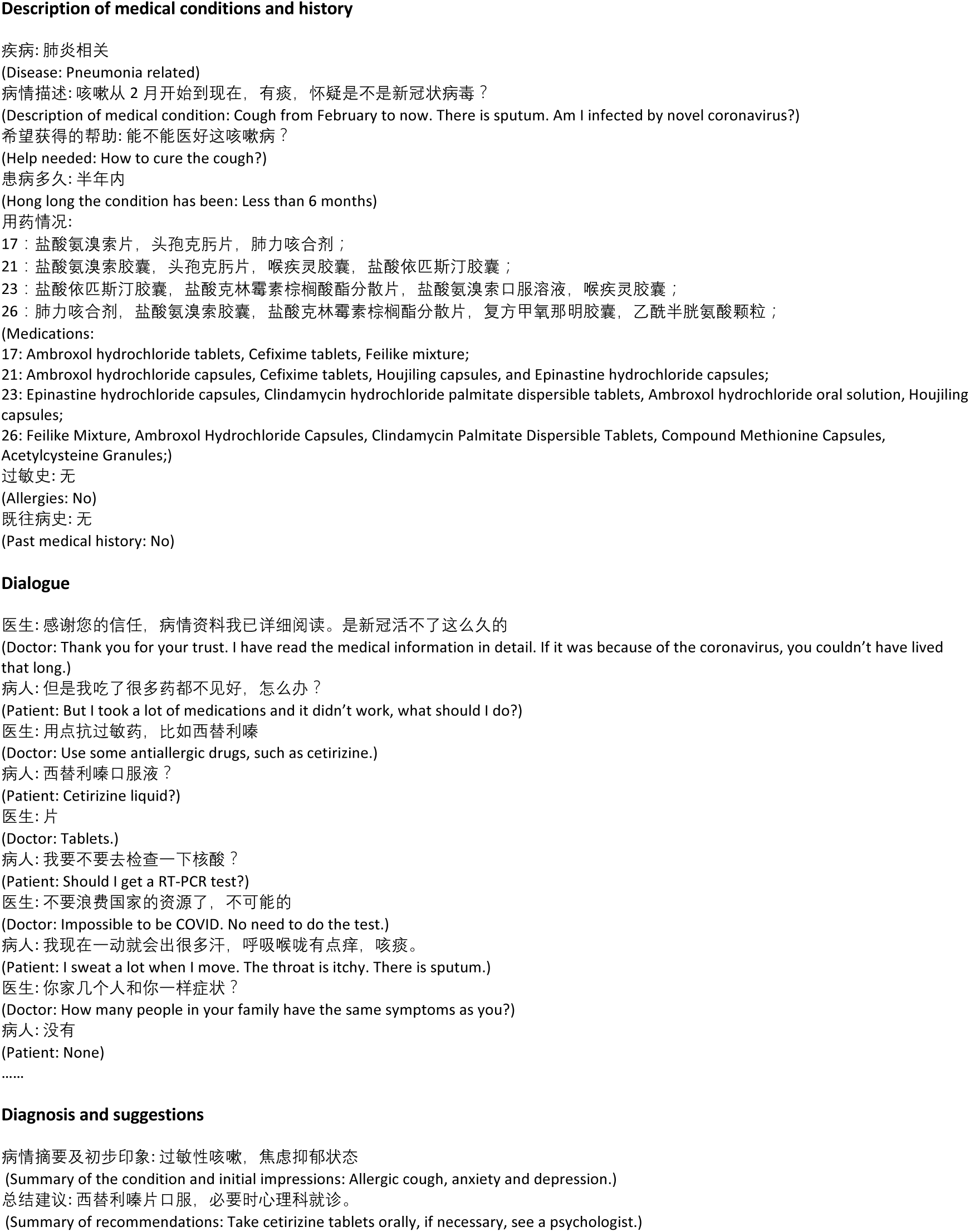
An exemplar consultation in the CovidDialog-Chinese dataset. It consists of (1) description of medical conditions and history of the patient, (2) dialogue between doctor and patient, and (3) diagnosis and treatment suggestions given by the doctor.

## 3. Methods

In this section, we present several well-established and state-of-the-art methods for dialogue generation. Given a dialogue containing a sequence of alternating utterances between patient and doctor, we process it into a set of pairs {(*s_i_*,*t_i_*)] where the target *t_i_* is a response from the doctor and the source *s_i_* is the concatenation of all utterances (from both patient and doctor) before *t_i_*. A dialogue generation model takes *s* as input and generates *t*. The size of the CovidDialog datasets is small. Directly training neural models on these small datasets would result in poor generalization on unseen data. To solve this problem, we utilize transfer learning, which pretrains the neural models on large corpus, then finetunes the pretrained models on the CovidDialog datasets.

### 3.1. Transformer

Generating response *t* from the conversation history *s* is a typical sequence-to-sequence (seq2seq) (Sutskever et al., 2014) modeling problem. Transformer (Vaswani et al., 2017) is an encoder-decoder architecture for sequence-to-sequence (seq2seq) modeling. Different from seq2seq models (Sutskever et al., 2014) that are based on recurrent neural networks (e.g., LSTM (Hochreiter and Schmidhuber, 1997), GRU (Chung et al., 2014)) which model a sequence of tokens via a recurrent manner and hence is computationally inefficient. Transformer eschews recurrent computation and instead uses self-attention which not only can capture the dependency between tokens but also is amenable for parallel computation with high efficiency. Self-attention calculates the correlation among every pair of tokens and uses these correlation scores to create “attentive” representations by taking weighted summation of tokens’ embeddings. Transformer is composed of a stack of building blocks, each consisting of a self-attention layer and a position-wise feed-forward layer. Residual connection (He et al., 2016) is applied around each of the two sub-layers, followed by layer normalization (Ba et al., 2016). Given the input sequence, an encoder, which is a stack of such building blocks, is applied to obtain a representation for each token. Then the decoder takes these representations as inputs and decodes the sequence of output tokens. To decode the *i*-th token, the decoder first uses self-attention to encode the already decoded sequence *y*_1_, ┅ *,y_i_*_−1_, then performs input-output attention between the encodings of *y*_1_, ┅*,y_i_*_−1_ and those of the input sequence. The “attentive” representations are then fed into a feed-forward layer. The three steps are repeated for multiple times. Finally, the representation is fed into a linear layer to predict the next token. The weight parameters in Transformer is learned by maximizing the conditional likelihood of output sequences conditioned on the corresponding input sequences.

### 3.2. GPT

The GPT model (Radford et al., a) is a language model (LM) based on Transformer. Different from Transformer which defines a conditional probability on an output sequence given an input sequence, GPT defines a marginal probability on a single sequence. Given a sequence of tokens *x*_1_, ┅*,x_n_*, an LM defines a probability on the sequence:

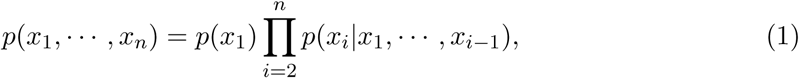

which basically predicts the next token based on the historical sequence. In GPT, *p*(*x_i_*|*x*_1_, ┅,*x_i_*_−1_) is defined using the Transformer decoder, which first uses a stack of self-attention and feed-forward layers (each followed by layer normalization) to encode *x*_1_, ┅, *x_i_*_−1_, then predicts *x_i_* from the encodings of *x*_1_, ┅, *x_i_*_−1_. The weight parameters are learned by maximizing the likelihood on the sequence of tokens. GPT-2 (Radford et al., b) is an extension of GPT, which modifies GPT by moving layer normalization to the input of each sub-block and adding an additional layer normalization after the final self-attention block. Byte pair encoding (BPE) (Sennrich et al., 2015) is used to represent the input sequence of tokens.

#### Pretrained GPT models for dialogue generation

DialoGPT (Zhang et al., 2019) is a GPT-2 model pretrained on English Reddit dialogues. The dataset is extracted from comment chains in Reddit from 2005 till 2017, comprising 147,116,725 dialogue instances with 1.8 billion tokens. Given a dialogue history *S* and a ground-truth response *T* = *x*_1_, ┅,*x_n_*, DialoGPT is trained to maximize the following probability

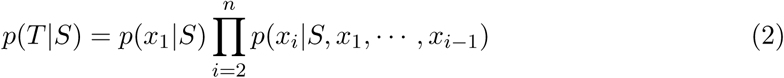

where the conditional probabilities are defined by the Transformer decoder. A maximum mutual information (MMI) (Li et al., 2015) scoring function is used to penalize generated responses that are bland. We finetune DialoGPT on our CovidDialog-English dataset for generating English COVID-19 dialogues. GPT2-chitchat^5^ is a GPT-2 model pretrained on Chinese Chatbot Corpus^6^ which contains about 14M dialogues and 500k-Chinese-Dialog^7^ which contains 500K Chinese dialogues. The training strategy of GPT2-chitchat is the same as that of DialoGPT. We finetune GPT2-chitchat on our CovidDialog-Chinese dataset for generating Chinese COVID-19 dialogues.

### 3.3. BERT-GPT

BERT-GPT (Wu et al., 2019) is a model used for dialogue generation where pretrained BERT is used to encode the conversation history and GPT is used to generate the responses. While GPT focuses on learning a Transformer decoder for text generation purposes, BERT (Devlin et al., 2018) aims to learn a Transformer encoder for representing texts. BERT’s model architecture is a multi-layer bidirectional Transformer encoder. In BERT, the Transformer uses bidirectional self-attention, whereas in GPT every token can only attend to context to its left. To train the encoder, BERT masks some percentage of the input tokens at random, and then predict those masked tokens by feeding the final hidden vectors (produced by the encoder) corresponding to the mask tokens into an output softmax over the vocabulary. Since BERT leverages context to both the left and the right for representing a token, it presumably has better representation power than GPT which only leverages context to the left. In dialogue generation, for the given conversation history, instead of using GPT for obtaining the representation, we can use a more powerful pretrained BERT to encode it. The BERT encoding of the conversation history is fed into GPT to generate the response.

In BERT-GPT, the pretraining of the BERT encoder and the GPT decoder is conducted separately, which may lead to inferior performance. Auto-Regressive Transformers (BART) (Lewis et al., 2019) has a similar architecture as BERT-GPT, but trains the BERT encoder and GPT decoder jointly. To pretrain the BART weights, the input text is corrupted randomly, such as token masking, token deletion, text infilling, etc., then the network is learned to reconstruct the original text. BART is pretrained on the data used in (Liu et al., 2019), consisting of 160Gb of news, books, stories, and web texts.

#### Pretrained BERT-GPT models for dialogue generation

BERT-GPT-Chinese (Wu et al., 2019) is a BERT-GPT model pretrained on Chinese corpus. For the BERT encoder in BERT-GPT-Chinese, it is set to the Chinese BERT (Cui et al., 2019), which is a large-scale pretrained BERT language model on Chinese texts. For the GPT decoder in BERT-GPT, it has the same architecture as BERT but applies lower-triangular mask for autoregressive text generation. The decoder is initialized with Chinese BERT’s weights. Then the decoder is pretrained with a maximum likelihood estimation (MLE) objective on a large-scale multidomain Chinese corpus. The resulting model consists of a bidirectional Transformer as the encoder, a unidirectional Transformer as the decoder, and an attention mechanism to connect them. The Chinese corpus used for pretraining is collected from the Large Scale Chinese Corpus for NLP^8^, including the following datasets: Chinese Wikipedia which contains 104M articles, News which contains 2.5 million news articles from 63,000 sources, Baike QA which is a wiki question answering (QA) dataset with 1.5 million QA pairs from 493 different domains, and Community QA which contains 4.1 million comments and 28 thousand topics. The total size of these datasets is 15.4 GB. We finetune BERT-GPT-Chinese on the CovidDialog-Chinese dataset for Chinese COVID-19 dialogue generation. For English COVID-19 dialogue generation, we finetune the pretrained BART model on the CovidDialog-English dataset.

## 4. Experiments

### 4.1. Experiments on the English Dataset

#### 4.1.1. Experimental Settings

For the English dataset, we split it into a training, a validation, and a test set based on dialogues, with a ratio of 8:1:1. Table 2 shows the statistics of the data split. The size of the vocabulary (number of unique English words) was set to x. The hyperparameters were tuned on the validation dataset. For all methods, we used the Adam (Kingma and Ba, 2014) optimizer with linear learning rate scheduling, setting the initial learning rate as 4e-5 and the batch size as 4. The objective is the cross entropy loss with label smoothing where the factor was set to 0.1. For pretrained models, we finetune them on the CovidDialog-English dataset for 5 epochs, while for the un-pretrained Transformer, we train it for 50 epochs. We set a checkpoint at the end of every epoch and finally take the one with the lowest perplexity on validation set as the final model. In response generation, for all models, we use beam search with beam width of 10 as our decoding strategy. For DialoGPT (Zhang et al., 2019), we used three variants with different sizes: DialoGPT-small, DialoGPT-medium, DialoGPT-large, with 117M, 345M and 762M weight parameters respectively. Maximum mutual information was not used.

**Table 2:**
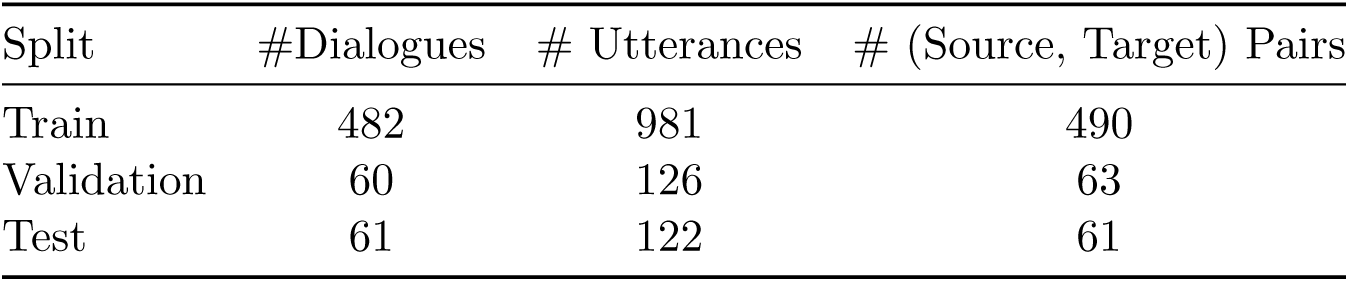
English dataset split statistics

| Split | #Dialogues | # Utterances | # (Source, Target) Pairs |
| --- | --- | --- | --- |
| Train | 482 | 981 | 490 |
| Validation | 60 | 126 | 63 |
| Test | 61 | 122 | 61 |

We performed automatic evaluation, using metrics including perplexity, NIST-*n* (Doddington, 2002) (where *n* = 4), BLEU-*n* (Papineni et al., 2002) (where *n* = 2 and 4), METEOR (Lavie and Agarwal, 2007), Entropy-*n* (Zhang et al., 2018) (where *n* = 4), and Dist-*n* (Li et al., 2015) (where *n* = 1 and 2). BLEU, METEOR, and NIST are common metrics for evaluating machine translation. They compare the similarity between generated responses and the ground-truth by matching *n*-grams. NIST is a variant of BLEU, which weights *n*-gram matches using information gain to penalize uninformative *n*-grams. Perplexity is used to measure the quality and smoothness of generated responses. Entropy and Dist are used to measure the lexical diversity of generated responses. For perplexity, the lower, the better. For other metrics, the higher, the better.

#### 4.1.2. Results

Table 3 summarizes the results achieved by different methods. From this table, we make the following observations. First, pretrained models including DialoGPT and BART in general perform better than un-pretrained Transformer. This demonstrates the effectiveness of transfer learning, which leverages external large-scale data to learn powerful representations of texts. Second, BART achieves lower perplexity than DialoGPT models. This is probably because BART is pretrained on a much larger and more diverse corpus than DialoGPT, which enables BART to better model the language. Third, DialoGPT-large performs better than BART on machine translation metrics including NIST, BLEU, and METEOR. This is probably because DialoGPT-large is pretrained on dialogue data and therefore tends to generate *n*-grams that are more related to dialogues. Fourth, on diversity-related metrics including Entropy and Dist, BART are on par with DialoGPT models. Note that the comparison between different architectures is not totally fair since they are pretrained on different corpus. Due to the lack of computing resources, we are not able to make a fair comparison by training these architectures on the same corpus. We will leave such a study to the future. The average length of the generated responses by different methods is close to that of the ground-truth, which is around 50.

**Table 3:** Performance on the CovidDialog-English test set.

|  | Transformer | DialoGPT-small | DialoGPT-medium | DialoGPT-large | BART |
| --- | --- | --- | --- | --- | --- |
| Perplexity | 263.1 | 28.3 | 17.5 | 18.9 | **15.3** |
| NIST-4 | 0.71 | 1.90 | 2.01 | **2.29** | 1.88 |
| BLEU-2 | 7.3% | 9.6% | 9.4% | **11.5%** | 8.9% |
| BLEU-4 | 5.2% | 6.1% | 6.0% | **7.6%** | 6.0% |
| METEOR | 5.6% | 9.0% | 9.5% | **11.0%** | 10.3% |
| Entropy-4 | 5.0 | 6.0 | **6.6** | **6.6** | 6.5 |
| Dist-1 | 3.7% | 9.5% | 16.6% | 13.9% | **16.8%** |
| Dist-2 | 6.4% | 22.9% | **36.7%** | 31.0% | **35.7%** |
| Avg. Len | 40.0 | 51.3 | 50.1 | 54.4 | 45.4 |

Figure 3 shows an example of generating a doctor’s response given the utterance of a patient. As can be seen, the response generated by BART is more relevant, informative, and human-like, compared with those generated by other baselines. BART’s response suggests the patient to get tested for COVID-19 since the patient stated that “I have all the symptoms except fever”. This response gives correct and informative medical advice: “get tested if you have fever, cough, or shortness of breath”, “if you are a smoker or have been in contact with someone with covid, get tested”. The response is human-like, with correct grammar and semantics. It begins with a welcome opening, then provides medical advice, and finally offers to further discuss via video. In contrast, the response generated by DialoGPT-large is not informative. It does not provide any useful medical advice. The response generated by DialoGPT-medium is informative, but not very relevant. The patient has no fever, but this response focuses on talking about the causes of fever. Similar to DialoGPT-large, the responses generated by DialoGPT-small and Transformer are uninformative.

**Figure 3:**
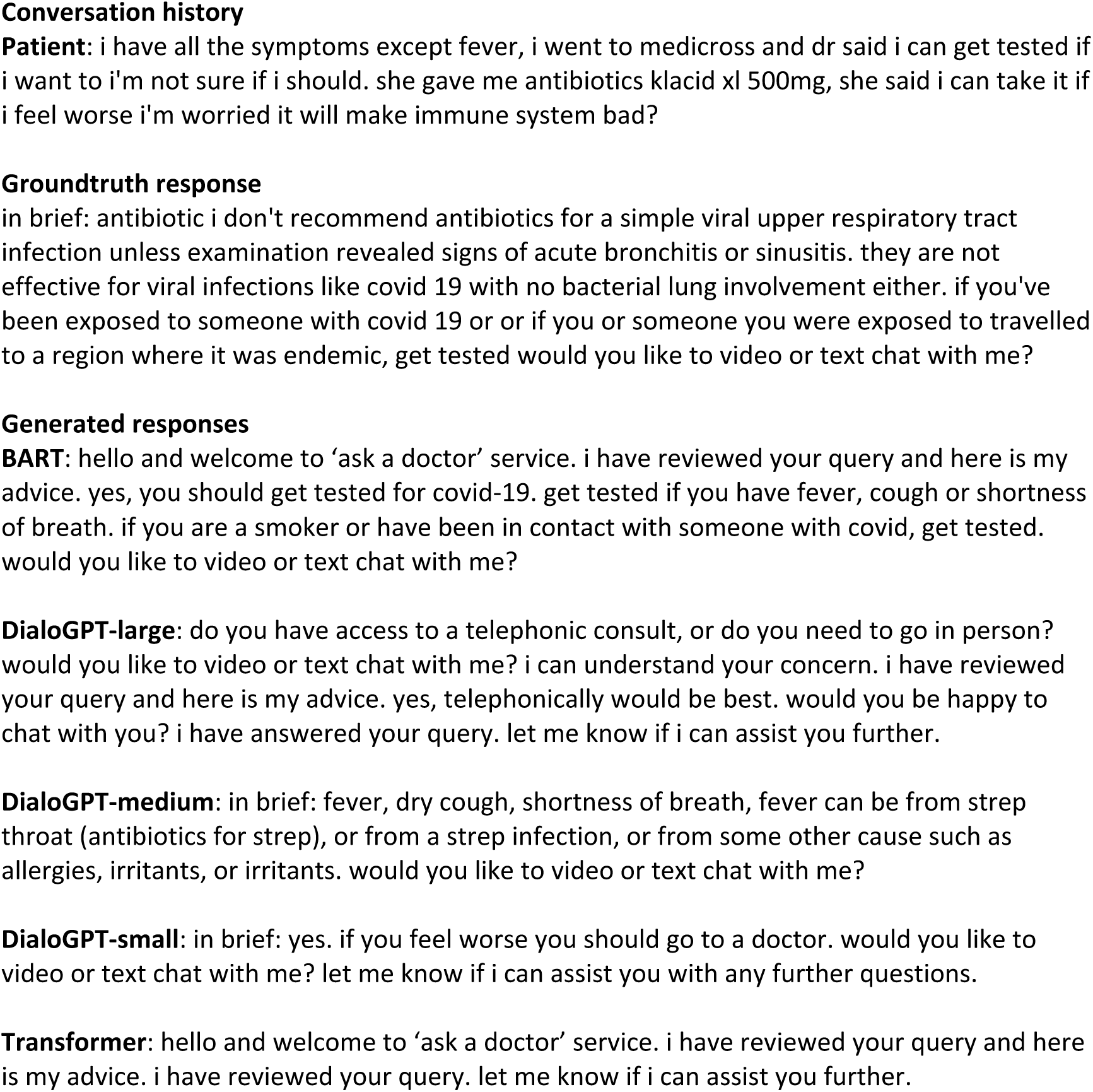
An example of generated English responses.

### 4.2. Experiments on the Chinese Dataset

#### 4.2.1. Experimental settings

Based on dialogues, we split the Chinese dataset into a training set, validation set, and test set, with a ratio of 8:1:1. Table 4 shows the statistics of the data split. The vocabulary size (number of unique Chinese characters) was set to 13317. The hyperparameters were tuned on the validation set. We stop the training procedure when the validation loss stops to decrease. For DialoGPT, we used the DialoGPT-small architecture where the number of layers in the Transformer was set to 10. The context size was set to 300. The embedding size was set to 768. The number of heads in multi-head self-attention was set to 12. The epsilon parameter in layer normalization was set to 1e-5. Network weights were optimized with Adam, with an initial learning rate of 1.5e-4 and a batch size of 8. The Noam learning rate scheduler with 2000 warm-up steps was used. In the finetuning of BERT-GPT, the max length of the source sequence and target sequence was set to 400. The encoder and decoder structures are similar to those in BERT, which is a Transformer with 12 layers and the size of the hidden states is 768. The network weights are optimized with stochastic gradient descent with a learning rate of 1e-4. For Transformer, we used the HuggingFace implementation^9^ and followed their default hyperparameter settings. During decoding for all methods, beam search with *k* = 50 was used. We evaluated the models using perplexity, NIST-4, BLEU-2, 4, METEOR, Entropy-4, and Dist-1, 2.

**Table 4:** Chinese dataset split statistics

| Split | # Dialogues | # Utterances | # (Source, Target) Pairs |
| --- | --- | --- | --- |
| Train | 870 | 7844 | 3922 |
| Validation | 109 | 734 | 367 |
| Test | 109 | 916 | 458 |

### 4.3. Results on the Chinese Dataset

Table 5 summarizes the results. From this table, we make the following observations. First, pretrained models including DialoGPT and BERT-GPT achieve lower perplexity than Transformer. This further demonstrates the effectiveness of transfer learning. Second, DialoGPT-MMI achieves better scores on machine translation metrics, which is consistent with the results on the CovidDialog-English dataset. Third, BERT-GPT achieves much better Dist scores than other methods. We manually checked the generated responses by BERT-GPT. Indeed, they are more diverse than others. Fourth, maximum mutual information (MMI) does not have a clear efficacy in improving the quality of generated responses.

**Table 5:** Performance on the CovidDialog-Chinese test set.

|  | Transformer | DialoGPT, no MMI | DialoGPT, with MMI | BERT-GPT |
| --- | --- | --- | --- | --- |
| Perplexity | 53.3 | 37.0 | 42.1 | **10.2** |
| NIST-4 | 0.39 | 0.41 | **0.45** | 0.30 |
| BLEU-2 | 5.7% | 5.5% | **6.2%** | 4.5% |
| BLEU-4 | 4.0% | 3.1% | **4.5%** | 2.7% |
| METEOR | **13.5%** | 13.4% | 13.3% | 11.1% |
| Entropy-4 | 7.9 | **9.4** | **9.4** | 8.3 |
| Dist-1 | 5.5% | 2.2% | 1.6% | **8.7%** |
| Dist-2 | 29.0% | 25.3% | 29.3% | **40.5%** |
| Avg Len | 37.5 | 43.3 | 55.6 | 18.7 |

Figure 4 shows an example of generating a doctor’s response given the utterance of a patient. The response generated by BERT-GPT matches with the ground-truth, both of which indicate that the patient has low risk of being infected. The responses generated by other methods are not understandable.

**Figure 4:**
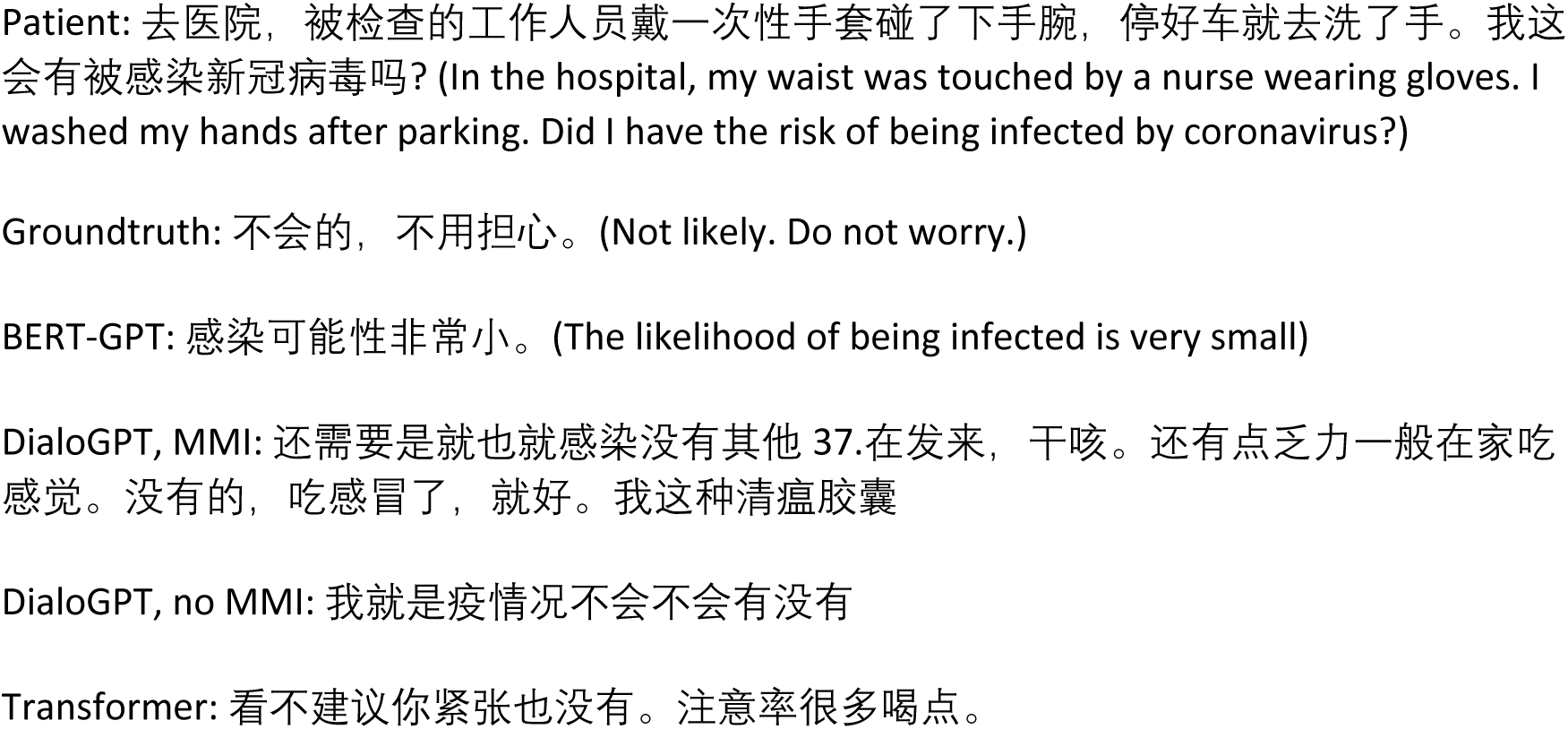
An example of generated Chinese responses.

## 5. Related Works

Many works have been devoted to developing medical dialogue systems. Please refer to (Laranjo et al., 2018) for a comprehensive review. Some methods (Lucas et al., 2017; Philip et al., 2017; Tanaka et al., 2017) predefine a sequence of steps or states which are used to guide the conversation. Other methods (Rhee et al., 2014; Ireland et al., 2016; Fitzpatrick et al., 2017) use predetermined templates to extract information from the conversation history and use rules to generate responses from the filled slots in the templates. These methods rely heavily on knowledge engineering and are difficult to be quickly adapted to a new and time-sensitive task such as COVID-19 dialogue generation.

Data-driven medical dialogue generation based on neural networks has been investigated in several works. Wei et al. (Wei et al., 2018) proposed a task-oriented dialogue system to make medical diagnosis automatically based on reinforcement learning. The system converses with patients to collect additional symptoms beyond their self-reports. Xu et al. (Xu et al., 2019) proposed a knowledge-routed relational dialogue system that incorporates medical knowledge graph into topic transition in dialogue management. Xia et al. (Xia et al.) developed a reinforcement learning (RL) based dialogue system for automatic diagnosis. They proposed a policy gradient framework based on the generative adversarial network to optimize the RL model. In these works, the neural models are trained from scratch on small-sized medical dialogue datasets, which are prone to overfitting.

## 6. Conclusions

In this work, we make the first attempt to develop dialogue systems that can provide medical consultations about COVID-19. To achieve this goal, we first collected two datasets – CovidDialog – which contain medical conversations between patients and doctors about COVID-19. Then on these datasets, we train dialogue generation models based on pretrained Transformer, DialoGPT, and BERT-GPT on large-scale dialogue datasets and other corpus. Experimental results show that these trained models are promising in generating clinically meaningful and linguistically high-quality consultations for COVID-19.

## Data Availability

COVID-Dialogue-Dataset-English is an English medical dialogue dataset about COVID-19 and other types of pneumonia. Patients who are concerned that they may be infected by COVID-19 or other pneumonia consult doctors and doctors provide advice. There are 603 consultations. COVID-Dialogue-Dataset-Chinese is a Chinese medical dialogue dataset about COVID-19 and other types of pneumonia. Patients who are concerned that they may be infected by COVID-19 or other pneumonia consult doctors and doctors provide advice. There are 1393 consultations.

https://github.com/UCSD-AI4H/COVID-Dialogue

## Footnotes

1. https://www.icliniq.com/en_US/

2. https://www.healthcaremagic.com/

3. https://www.healthtap.com/

4. https://www.haodf.com/

5. https://github.com/yangjianxin1/GPT2-chitchat

6. https://github.com/codemayq/chinese_chatbot_corpus

7. https://drive.google.com/file/d/1nEuew_KNpTMbyy7BO4c8bXMXN351RCPp/view

8. https://github.com/brightmart/nlp_chinese_corpus

9. https://github.com/huggingface/transformers

